# Spotting the Silent Threat: Early Detection of Subclinical Tuberculosis in High-Risk Individuals

**DOI:** 10.1101/2025.02.24.25322763

**Authors:** Evangeline Ann Daniel, Manohar Nesakumar, S Hemalatha, Nancy Hilda, Kannan Thiruvengadam, Umashankar Vetrivel, Anbalagan Selvaraj, Sathyamurthi Pattabiraman, Brindha Bhanu, Amsaveni Sivaprakasam, Sudhakar Natarajan, Vandana Kulkarni, Rajesh Karyakarte, Mandar Paradkar, Shri Vijay Bala Yogendra Shivakumar, Vidya Mave, Padmapriyadarsini Chandrasekaran, Amita Gupta, Luke Elizabeth Hanna

**Author notes:** **Corresponding author** Dr Luke Elizabeth Hanna, Scientist F & Head, Department of Virology and Biotechnology, ICMR-National Institute for Research in Tuberculosis, No 1, Mayor Sathiyamoorthy Road, Chetpet, Chennai – 600 031, India.

## Abstract

The dynamic spectrum of TB often results in underdiagnosis warranting the need for better diagnostics to accurately detect *Mtb* in diagnostically challenging cohorts. Household contacts of newly diagnosed TB patients who developed TB (Progressors) and those who remained healthy (Non-progressors) during a two-year follow-up cohort study were included. *Mtb* ccfDNA was detected in the plasma by targeting insertion sequences IS6110 and IS1081 using ddPCR. The assay yielded a sensitivity of 90·9% in detecting subclinical TB cases and 81·8% in possible TB cases. Further, the test detected *Mtb* ccfDNA in progressors even at six months prior to TB diagnosis at a sensitivity of 79·0%. In about 55·0% and 50·0% of cases *Mtb* ccfDNA could be detected as early as 12 months and 18 months prior to development of active disease, respectively. The test demonstrated excellent sensitivity (100·0%) in detecting extra-pulmonary TB cases up to six months prior to TB disease development.

## INTRODUCTION

The “End TB strategy” of the World Health Organization envisions the reduction of TB incidence by 80% and TB deaths by 90% by 2030 ^1^. The key strategy for achieving this goal is the systematic screening of individuals with high risk to identify those who will progress to active TB disease and providing appropriate intervention to those with higher susceptibility^2^. Contrary to the long-established notion of TB manifesting either in the asymptomatic latent form or the symptomatic active disease, it has become evident now that TB infection is a dynamic continuum. The spectrum includes two additional clinical states - incipient TB, where *Mtb* is slowly replicating but does not exhibit any detectable abnormalities; and subclinical TB, where *Mtb* is actively replicating and could be detected by radiological and microbiological tests, but with either mild/unrecognizable or no clinical symptoms^3^. This pool of individuals with subclinical TB may remain infectious in the community for years without exhibiting symptoms, and therefore represent a critical pool for TB transmission^4^. This serious state calls for prioritizing active case finding using non-symptom based, point-of-care tests with superior performance characteristics. Despite technological advances, the major challenge in devising such a test is the low bacillary load in subclinical TB ^5^.

Detection of pathogen-derived cell free DNA has been gaining much attention in recent years for the diagnosis of several clinical conditions. Microbial DNA fragments, derived from pathogens or dying host cells or tissues, are released into the bloodstream and remain in circulation for a number of days ^6^. Since 70% of cfDNA in the plasma are <300bp, cfDNA can easily cross the kidney barrier and can also be detected in urine. Several recent studies have harnessed the highly sensitive digital droplet PCR (ddPCR) to detect ccfDNA with substantial sensitivity and specificity. A recent meta-analysis showed that ddPCR targeting combination of two *Mtb* targets IS6110 and IS1081 gave an AUC of 0·9^7^.

The insertion sequence IS6110 is a mobile transposable element found solely in the *Mtb* complex and is widely used as a target for TB diagnosis ^8^. Multiple copies of IS6110 have been identified at different genomic locations *in Mtb* (up to 25 per genome) ^9^. IS1081 is a relatively stable element with 5-7 copies per genome ^10^. The combined use of the two insertion sequences has been reported to substantially improve the sensitivity of the ddPCR assay ^11,12^.

The ability of ddPCR to detect very low copy numbers of the target and its potential to detect *Mtb-*derived ccfDNA prompted us to explore its utility in detecting *Mtb-*ccfDNA in latently infected individuals who subsequently progressed to active TB (Progressors), including individuals with subclinical TB, with an assumption that if the test is able to detect *Mtb* DNA in Progressors much earlier than the development of active infectious TB disease, the implications of the finding would have far reaching benefits for TB control and elimination.

## METHODS

### Ethical statement

The parent study protocol received ethical approval from the Institutional Ethics Committees of the ICMR-National Institute for Research in Tuberculosis (ICMR-NIRT) in Chennai, India, Byramjee Jeejeebhoy Government Medical College (BJGMC) in Pune, India, and Johns Hopkins University (JHU) in Baltimore, USA.

### Study cohort

Our study population included a sub-cohort from a cohort of healthy household contacts (HHCs) of TB patients who were longitudinally followed up for two years from August 2014 to December 2017 as part of a cohort study called the Cohort for Tuberculosis Research by the Indo-US Medical Partnership (CTRIUMPh) study carried out at the ICMR-National Institute for Research in Tuberculosis (NIRT) and Byramjee Jeejeebhoy Government Medical College (BJGMC), Pune as well as from other assorted studies of ICMR-NIRT carried out during the same period^13^. HHCs were defined as all adults and children living in the same house as an adult (>18yrs) with active PTB. Specimens collected include sputum for AFB, Xpert MTB/Rif, solid and liquid TB cultures; whole blood for TST, IGRA, HIV, and HbA1c, and chest radiographs. Further, clinically indicated TB diagnostics were carried out to identify tuberculosis in suspected extrapulmonary sites, such as lymph node biopsies for lymphadenopathy, pleural fluid examinations for pleural effusion, and abdominal ultrasounds for abdominal pain. All HHCs underwent thorough clinical assessment at baseline, as well as at 4-6 months, 12 months and 24 months to rule out TB. HHCs who developed active TB during follow-up were categorized as Progressors and included in the present analysis. Table 1 provides the definition of the different categories of Progressors classified based on their outcome. HHCs who did not progress to active TB and remained healthy during the entire follow-up period were categorized as ‘Non-Progressors’. The participant selection flow diagram is provided in Supplementary Figure 1.

**Table 1.** Criteria for defining the outcome of the study participants.

| Outcome | Definition |
| --- | --- |
| **Confirmed incident PTB** | Appearance of symptoms/signs suggestive of PTB disease, with *Mtb* growth on culture. |
| **Subclinical PTB** | 1. *Mtb* DNA detected on Xpert MTB/Rif when culture was not performed or if culture was negative.<br>2. AFB detected on smear microscopy, if culture and Xpert MTB/Rif were not performed or if results were negative. |
| **Clinical/possible incident TB** | Appearance of symptoms suggestive of PTB disease but culture, Xpert MTB/Rif and AFB smear microscopy were not performed, or results are negative. |
| **Confirmed incident EPTB** | Appearance of signs/symptoms of any form of EPTB disease and with a positive culture for *Mtb* growth and/or positive Xpert MTB/Rif test on the EPTB specimen. |
| **Clinical/possible incident EPTB** | Appearance of signs/symptoms of any form of EPTB disease but without culture, Xpert MTB/Rif, AFB smear microscopy, and histopathology results, or with negative test results |

*PTB: Pulmonary Tuberculosis; EPTB: Extra pulmonary Tuberculosis; Mtb: Mycobacterium* *tuberculosis; Xpert MTB/Rif: Test for detection of Mtb and Rifampin resistance mutations;* *AFB: Acid fast bacilli*

### Extraction of circulating cell free DNA

ccfDNA was extracted from 1mL of plasma collected at baseline, month two/four, month six/twelve, and at the time of TB breakdown (as available) using the QIAamp MinElute ccfDNA Kit (Qiagen, Germany) according to the manufacturer’s protocol. Briefly, 1 mL of plasma was mixed with 30 μL of magnetic bead suspension, 55 μL of Proteinase K and 150 μL of bead binding buffer and incubated for 10 minutes at room temperature, while shaking end-over-end. The tube was then placed in a magnetic rack and the supernatant was discarded. After this, 200 μL of bead elution buffer was added to the pellet. The tube was positioned in the magnetic rack, and the supernatant was carefully transferred to a fresh tube. 300 μL of buffer ACB was added to the supernatant and mixed well. The mixture was then applied to the QIAamp UCP MinElute Column and centrifuged at 6000 x g for 1 minute. Following this, 500 μL of buffer ACW2 was added and centrifuged again at 6000 x g for 1 minute. The ccfDNA was eluted from the column by adding 30 μL of ultraclean water. The extracted DNA was stored at -80^°^C for further use.

### Detection of *Mtb*-specific multicopy insertion sequences, IS6110 and IS1081, using ddPCR

Primers and probes targeting the *Mtb* insertion sequences IS6110 and IS1081 were synthesized based on a previously published study ^12^. The sequence of the primers and probes used are provided in Supplementary Table 1.

The ddPCR platform includes an Automated Droplet Generator (autoDG), PX1 PCR Plate Sealer, C1000 Touch Thermal Cycler and the QX200 Droplet Reader, all manufactured by Bio-Rad (Hercules, CA, USA). The initial ddPCR reaction mixture consisted of the following components: 11 μL of 2X ddPCR Supermix for Probes (No dUTP), 0.88 μL of 10mM probe, 1 μL of 10mM each of forward and reverse primers, and DNA template. A reaction pool of slightly over 20μL (that is 22μL) was established to guarantee the transfer of precisely 20μL of the mixture into the DG32 automated droplet generator cartridge. The reaction plate was covered with a pierceable PCR plate heat seal foil (Bio-Rad, Hercules, CA, USA) and sealed using the PX1 PCR Plate Sealer at 180^°^C for 5 seconds. The sealed plate was loaded onto the autoDG for droplet generation. The new plate containing the generated droplets was again heat sealed. The sealed reaction plate containing ∼40μL reaction volume was transferred to the C1000 Touch Thermal Cycler for end point amplification.

The annealing temperature of the primers was first optimized using the CFX96 Touch Real-Time PCR Detection System (Bio-Rad, Hercules, CA, USA) and an annealing temperature of 56^°^C was found to be optimum for a temperature gradient of 54-60^°^C. The following cycling conditions were used for IS6110 and IS1081 amplification: initial denaturation at 95^°^C for 10 minutes, followed by 40 cycles of denaturation at 94^°^C for 30 seconds and annealing at 56^°^C for 1 minute, and a final step at 98^°^C for 10 minutes for signal stabilization. A ramp rate of 2°C per second was employed to ensure that every droplet reached the precise temperature at each stage of the cycle. The plate was incubated at 12^°^C for 4 hours and then at room temperature for 45 minutes prior to droplet reading in order to stabilize the droplets. The droplets were then read using the QX200 Droplet Reader to detect the number of positive droplets. The droplet reader uses the QuantaSoftTM software, version 1.7.4 (Bio-Rad, Hercules, CA, USA), to measure the fluorescence signal within each droplet. The software counts the number of positive and negative droplets and calculates the number of copies per microlitre (copies/μL) based on poisson statistics. Wells with fewer than 10,000 detected droplets were excluded from subsequent analysis. Inter-and-intra assay variability were ruled out by setting up the assay in duplicate and repeating the samples in different assays. The lower and upper limit of detection were determined using extracted H37Rv DNA as positive control. Starting with an initial concentration of 300ng/mL, the H37Rv DNA was diluted in nuclease free water to make serial 10-fold dilutions of the DNA ranging from 10^1^ (30ng) to 10^8^ (3fg). The 10^5^ and 10^8^ dilutions of H37Rv DNA was used as positive control in every assay.

### Statistical analysis

GraphPad Prism Version 10.1.1 (GraphPad software, MA, USA) was used to generate graphs and perform statistical analyses. Data are presented as mean ± standard deviation (SD). Significant differences in the mean IS6110 and IS1081 copy number between the groups as well as at various time points prior to TB breakdown in the Progressor group were evaluated using the Kruskal-Wallis test followed by Dunn’s multiple comparison test. Mann-Whitney test was used to evaluate significant differences in the mean IS6110 and IS1081 copy number between pulmonary and extra pulmonary TB cases. A *p-*value <0.05 was considered significant. Receiver Operating Characteristic (ROC) curve was constructed to determine the diagnostic accuracy of the ddPCR assay.

## RESULTS

### Characteristics of the study population

The clinical and demographic profile of the study participants is provided in Table 2. The participants were all healthy household contacts (HHCs) of index TB cases who were recruited along with the index cases and kept on longitudinal follow-up for a period of two years. Among the 3011 HHCs, we identified 46 HHCs who were asymptomatic at the time of enrolment but subsequently developed active TB disease. Among the 46 incident TB cases, 24 (52·2%) were categorized as confirmed incident TB, 11 (23·9%) as subclinical TB cases and 11 (23·9%) as possible incident TB. Twice the number (92) of HHCs who remained healthy throughout the entire follow-up (Non-progressors) were randomly selected and used as controls. The mean age of the 46 incident TB cases (18 males and 28 females) was 28·1 ± 1·9 years, while that of the 92 Non-progressors (31 males and 61 females) was 33·6 ± 1·5 years. Among the Progressors, 36 (76·6%) were tested positive for pulmonary TB (PTB) by either culture, smear or CBNAAT, and/or had radiological evidence of TB. Ten individuals (21·3%) were diagnosed with extrapulmonary TB (EPTB) by CBNAAT and/or clinical/radiological evidence of TB. Five (50%) of the 10 EPTB cases had pleural effusion, two (20%) had cold abscess, two (20%) had TB lymphadenitis and one (10%) had genitourinary TB. One progressor was diagnosed based on clinical/radiological evidence as having both PTB and EPTB. All 46 incident TB cases had stored plasma samples from at least 2 time points (baseline and at the time of TB breakdown). In addition, 13 individuals had plasma stored at 4 times points, and 3 had samples stored at 3 time points.

**Table 2.**
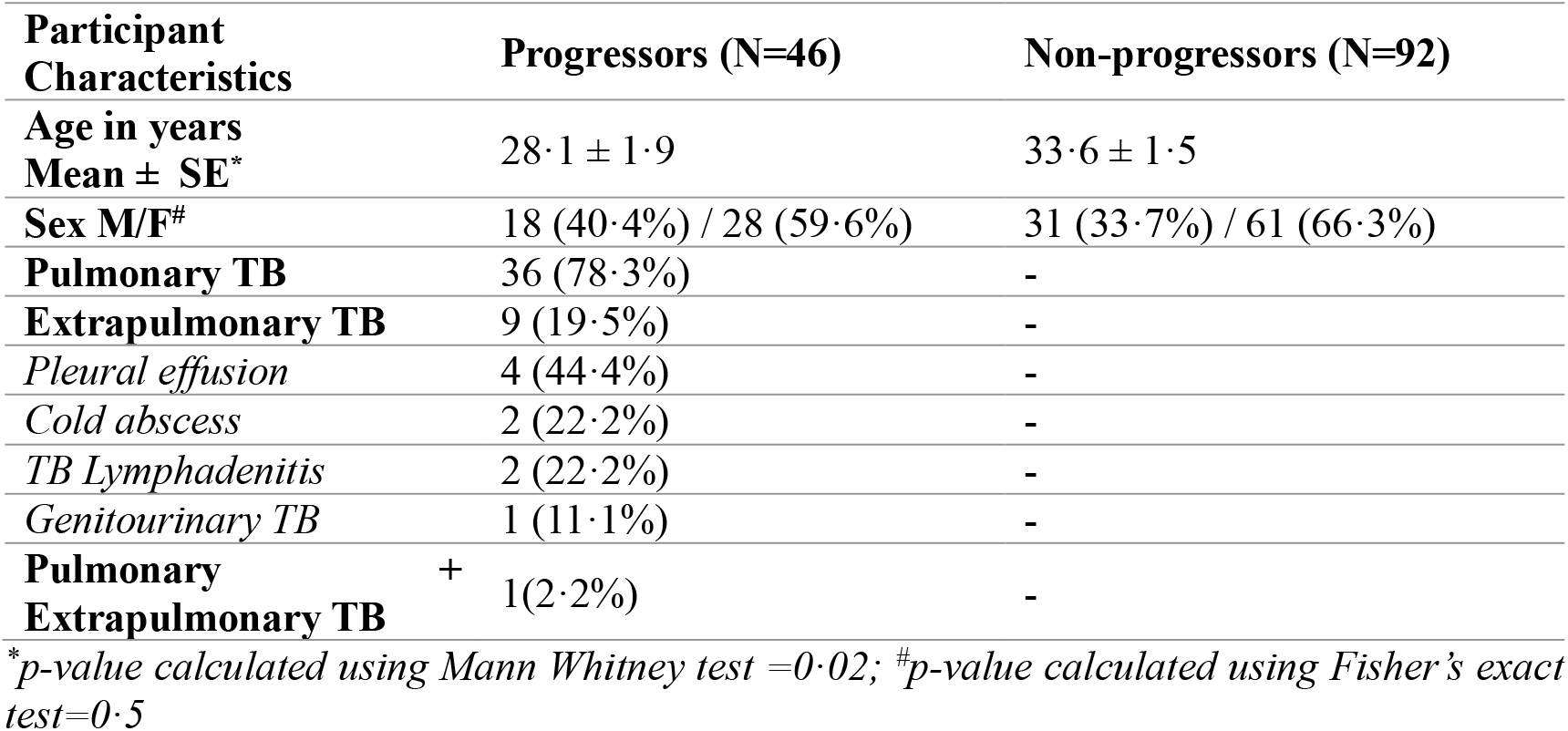
Clinical and demographic characteristics of the study participants.

### Detection of circulating cell free DNA in plasma samples

We determined the sensitivity of detection of ccf *Mtb* DNA using ddPCR in plasma samples collected at various longitudinal time points. At the time point of TB diagnosis, ddPCR could detect ccf DNA in 40/46 Progressors, giving a sensitivity of 87% (95% CI 74·3-93·9). Individually, ddPCR detected IS6110 in 38/46 (82·6%) Progressors and IS1081 in 23/46 (50·0%) Progressors. Two out of 92 Non-progressors (2.2%) tested positive for ccf DNA (IS1081) giving a specificity of 97·8% (95% CI 92·4-99·2). We tested these two individuals longitudinally, but did not detect *Mtb* DNA in any of the subsequent visit samples. A representative image of the amplitude plot and event count of the ddPCR assay is provided in Supplementary Figure 2.

Very interestingly, ccf DNA was detected in 15/19 Progressors up to 6 months prior to development of active TB disease, giving a sensitivity of 75% (95% CI 53·1-88·8). Going backwards the sensitivity declined gradually, but ccf DNA could be detected in 50% (10/20) of Progressors even as early as 18 months prior to the time of TB breakdown. Fig. 1A shows the mean copy number of the target detected in 1 mL of plasma in Progressors at various time points prior to TB breakdown. Table 3 shows the number of positives detected based on the two targets separately and together.

**Table 3.** Sensitivity of ddPCR assay in detecting *Mtb* ccfDNA in Progressors at various time points prior to TB breakdown.

| Time point of sample collection | No of samples | IS6110&IS1081 |  |  | IS6110 |  |  | IS1081 |  |  |
| --- | --- | --- | --- | --- | --- | --- | --- | --- | --- | --- |
|  |  | Positives detected (N) | Sensitivity (%) | Specificity (%) | Positives detected (N) | Sensitivity (%) | Specificity (%) | Positives detected (N) | Sensitivity (%) | Specificity (%) |
| At the time of TB breakdown | 46/46 | 40/46 | 87.0<br>(74.3-93.9) | 97.8<br>(92.4-99.6) | 38/46 | 82.6<br>(69.3-90.9) | 100.0<br>(96.0-100.0) | 23/46 | 50.0<br>(36.1-63.9) | 97.8<br>(92.4-99.6) |
| ≤6 months prior to TB breakdown | 20/46 | 15/19 | 79.0<br>(56.7-91.5) | 97.8<br>(92.4-99.6) | 12/19 | 63.2<br>(41.0-80.9) | 100.0<br>(96.0-100.0) | 5/19 | 26.3<br>(11.8-48.8) | 97.8<br>(92.4-99.6) |
| 7-11 months prior to TB breakdown | 15/46 | 9/15 | 60.0<br>(35.8-80.2) | 97.8<br>(92.4-99.6) | 8/15 | 53.3<br>(30.1-75.2) | 100.0<br>(96.0-100.0) | 4/15 | 26.7<br>(10.9-52.0) | 97.8<br>(92.4-99.6) |
| 12-18 months prior to TB breakdown | 20/46 | 11/20 | 55.0<br>(34.2-74.2) | 97.8<br>(92.4-99.6) | 8/20 | 40.0<br>(21.9-61.3) | 100.0<br>(96.0-100.0) | 7/20 | 35.0<br>(18.1-56.7) | 97.8<br>(92.4-99.6) |
| >18 months prior to TB breakdown | 20/46 | 10/20 | 50.0<br>(29.9-70.1) | 97.8<br>(92.4-99.6) | 8/20 | 40.0<br>(21.9-61.3) | 100.0<br>(96.0-100.0) | 4/20 | 20.0<br>(8.1-41.6) | 97.8<br>(92.4-99.6) |

**Fig. 1:**
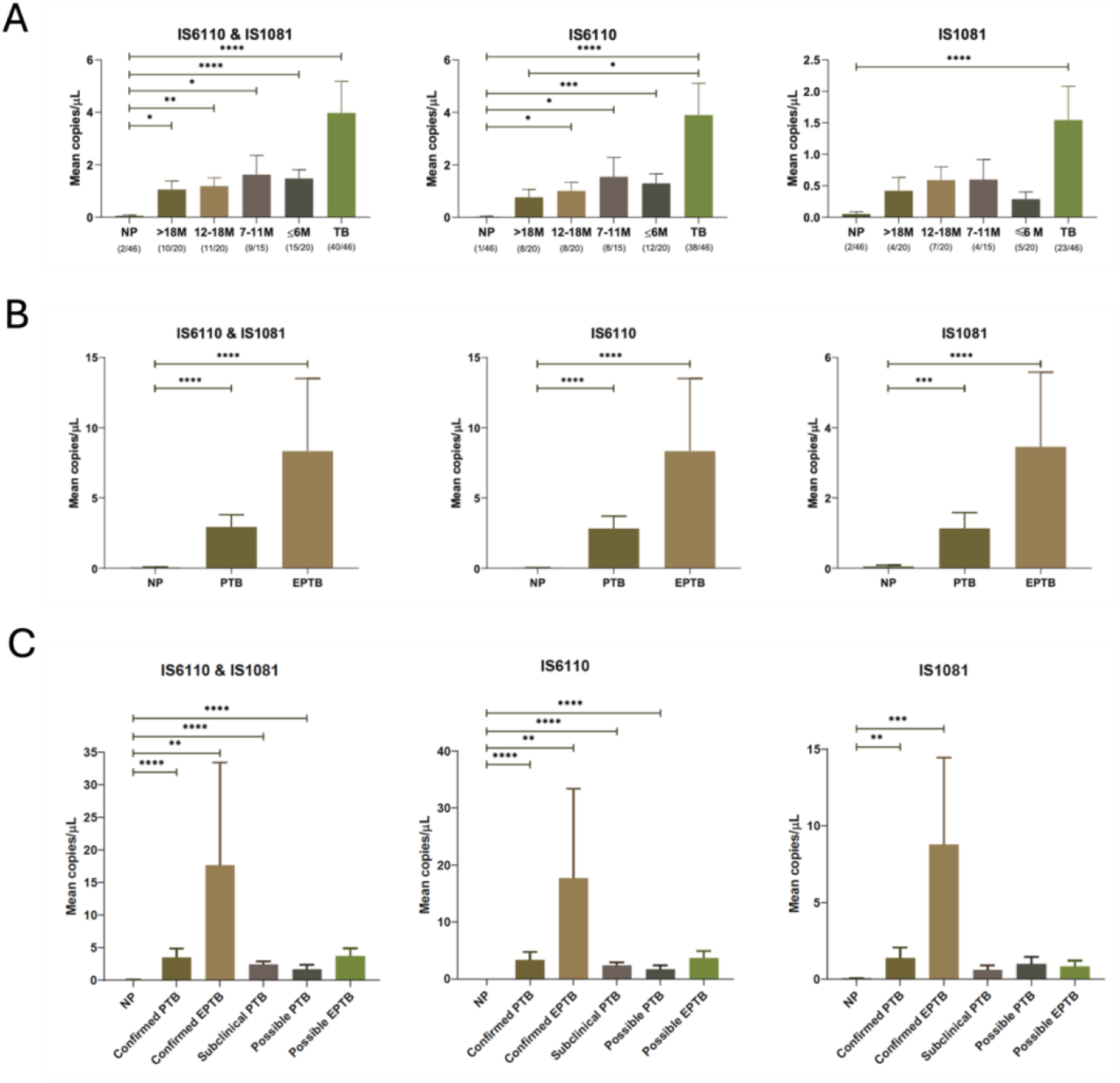
Mean copies of IS6110 and IS1081 detected in Progressors. (A) at various time points prior to TB breakdown (B) diagnosed with extra-pulmonary TB (C) with sub-clinical and possible TB *Significant differences in the mean IS6110 and IS1081 copy number between the groups as well as at various time points prior to TB breakdown in the Progressor group were evaluated using the Kruskal-Wallis test followed by Dunn’s multiple comparison test. A p-value <0*.*05 was considered significant* *PTB: Pulmonary Tuberculosis; EPTB: Extrapulmonary Tuberculosis*

### Higher sensitivity of ddPCR in detecting circulating cell-free *Mtb* DNA in progressors with extra pulmonary tuberculosis

Progressors diagnosed with extra pulmonary TB (EPTB) had significantly higher copies of IS6110 (8·3 ± 5·2) than those with pulmonary TB (PTB) (2·8 ± 0·9). Similarly, higher copy numbers of IS1081 (3·5 ± 2·1) were detected in EPTB patients who progressed to active TB disease as compared to pulmonary TB patients (1·1 ± 0·5). The sensitivity of detecting both IS6110 and IS1081 in EPTB progressors at the time of TB breakdown and up to 6 months prior to development of active TB was 100·0% (Fig. 1B; Table 4).

**Table 4.**
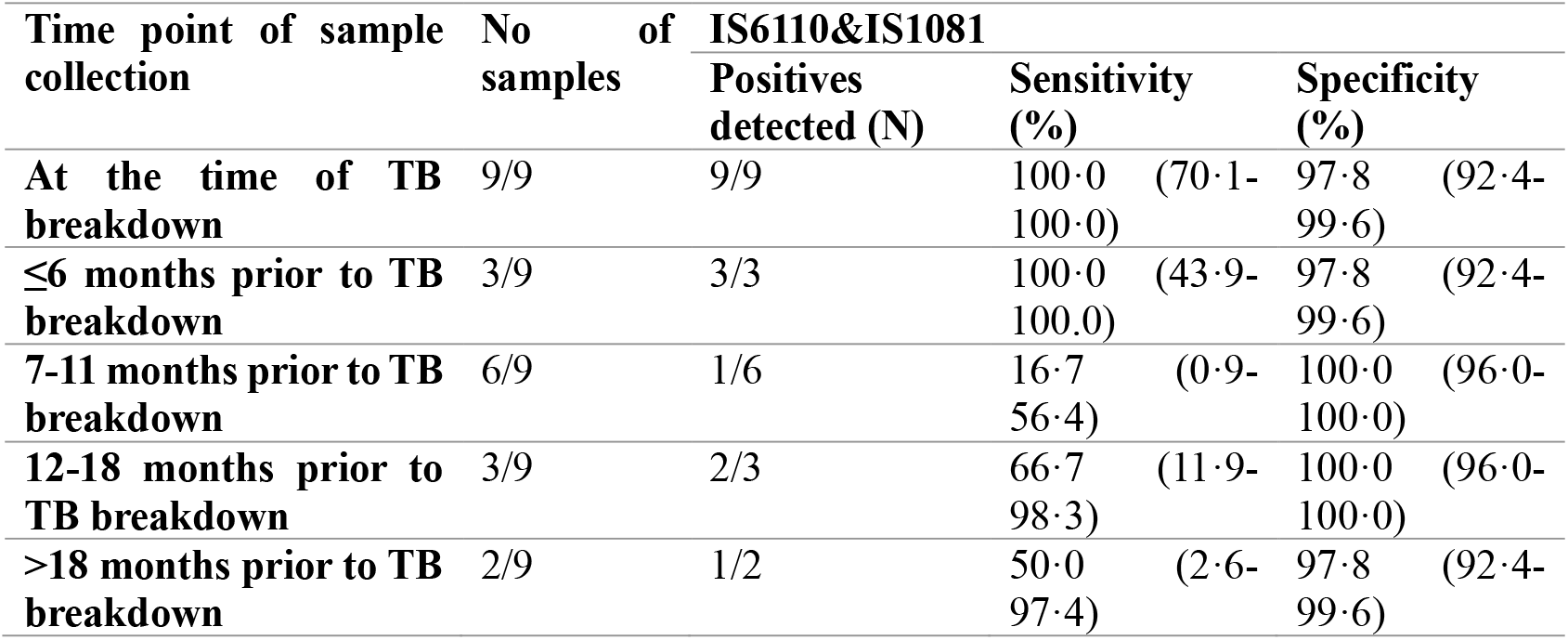
Sensitivity of ddPCR assay in detecting extra pulmonary TB cases at various time points prior to TB breakdown.

| Time point of sample collection | No of samples | IS6110&IS1081 |  |  |
| --- | --- | --- | --- | --- |
|  |  | Positives detected (N) | Sensitivity (%) | Specificity (%) |
| At the time of TB breakdown | 9/9 | 9/9 | 100.0 (70.1-100.0) | 97.8 (92.4-99.6) |
| ≤6 months prior to TB breakdown | 3/9 | 3/3 | 100.0 (43.9-100.0) | 97.8 (92.4-99.6) |
| 7-11 months prior to TB breakdown | 6/9 | 1/6 | 16.7 (0.9-56.4) | 100.0 (96.0-100.0) |
| 12-18 months prior to TB breakdown | 3/9 | 2/3 | 66.7 (11.9-98.3) | 100.0 (96.0-100.0) |
| >18 months prior to TB breakdown | 2/9 | 1/2 | 50.0 (2.6-97.4) | 97.8 (92.4-99.6) |

### Sensitivity of ddPCR in detecting circulating cell-free *Mtb* DNA in subclinical TB and possible TB cases

Out of the 11 subclinical TB cases, ccf DNA was detected in 10 individuals giving a sensitivity of 90.9%. Among the 11 possible TB cases who were diagnosed based only on clinical/radiological evidence, 9 tested positive for ccf DNA, giving a sensitivity of 81.8%. The mean target copy numbers/20μL in subclinical and possible TB cases is shown in Fig. 1C. The sensitivity, specificity and AUC of the ddPCR assay in detecting subclinical and possible PTB and EPTB cases are provided in Table 5.

**Table 5.** Sensitivity of ddPCR assay in detecting *Mtb* ccfDNA in subclinical and possible TB cases.

| Outcome | No of samples at the time of TB breakdown | IS6110&IS1081 |  |  | IS6110 |  |  | IS1081 |  |  |
| --- | --- | --- | --- | --- | --- | --- | --- | --- | --- | --- |
|  |  | Positives detected (N) | Sensitivity (%) | Specificity (%) | Positives detected (N) | Sensitivity (%) | Specificity (%) | Positives detected (N) | Sensitivity (%) | Specificity (%) |
| Confirmed PTB | 21 | 18/21 | 85.7<br>(65.4-95.0) | 97.8<br>(92.4-99.6) | 16/21 | 76.2<br>(54.9-89.4) | 100.0<br>(96.0-100.0) | 10/21 | 47.6<br>(28.3-67.6) | 97.8<br>(92.4-99.6) |
| Confirmed EPTB | 3 | 3/3 | 100.0<br>(43.9-100.0) | 98.9<br>(94.1-99.9) | 3/3 | 100.0<br>(43.9-100.0) | 100.0<br>(96.0-100.0) | 3/3 | 100.0<br>(43.9-100.0) | 98.9<br>(94.1-99.9) |
| Subclinical PTB | 11 | 10/11 | 90.9<br>(62.3-99.5) | 97.8<br>(92.4-99.6) | 10/11 | 90.9<br>(62.3-99.5) | 100.0<br>(96.0-100.0) | 4/11 | 36.4<br>(15.2-64.6) | 97.8<br>(92.4-99.6) |
| Possible PTB | 5 | 3/5 | 60.0<br>(23.1-92.9) | 98.9<br>(94.1-99.9) | 3/5 | 60.0<br>(23.1-92.9) | 100.0<br>(96.0-100.0) | 3/5 | 60.0<br>(23.1-92.9) | 97.8<br>(92.4-99.6) |
| Possible EPTB | 6 | 6/6 | 100.0<br>(61.0-100.0) | 97.8<br>(92.4-99.6) | 6/6 | 100.0<br>(61.0-100.0) | 100.0<br>(96.0-100.0) | 3/6 | 50.0<br>(18.8-81.2) | 97.8<br>(92.4-99.6) |

### Diagnostic performance of *Mtb* ccf DNA detection by IS6110-IS1081 ddPCR compared to routine diagnostic tests

We compared the sensitivity of the routine tuberculosis diagnostic tests with the concurrent detection of IS6110 and IS1081 by ddPCR. It was very encouraging to note that ccfDNA detection by ddPCR gave a diagnostic sensitivity of 87·0%, which was higher than that of all the currently available routine tests put together (76·1%). Table 6 provides the sensitivity of each of the routine diagnostic tests as well as ccfDNA detection by ddPCR.

**Table 6.** Sensitivity of ddPCR assay and routine diagnostic tests in detecting TB cases.

| Method | No. of positives detected (N) | Sensitivity |
| --- | --- | --- |
| Routine diagnostic tests (AFB smear, culture, NAAT) | 35/46 | 76.1% |
| Culture | 13/46 | 28.3% |
| AFB smear | 10/46 | 21.7% |
| NAAT | 23/46 | 50.0% |
| IS6110 & IS1081 ddPCR | 40/46 | 87.0% |

## DISCUSSION

Effective prevention and treatment of tuberculosis necessitates early diagnosis of individuals with subclinical/asymptomatic TB, underscoring the pivotal role of laboratories in swiftly and precisely identifying cases for appropriate management. Early detection of cases is hampered by the limitations of the currently available diagnostic tests. With a higher turnaround time, high risk of contamination and requirement of trained microbiologists, the gold standard *Mtb* culture test does not meet the requirements of a rapid test ^14^. Though quick and cost-effective, AFB smear does not differentiate viable from dead *Mtb*, and *Mtb* from *Mtb* complex ^15^. It also has a compromised sensitivity as it typically requires 5000-10000 bacilli per mL of sputum for a positive result ^16^. The widely endorsed Gene Xpert test requires a standard curve to determine the copy number. Evidence from the present study also reveals the low sensitivity of the routine TB diagnostics in detecting TB cases (Culture 27·7%; Smear 21·3% and NAAT 51·1%). Hence the search for non-sputum based, host/pathogen derived biomarker tests that are both rapid as well as sensitive continues.

There is evidence from existing literature that *Mtb*-ccfDNA circulates at very low concentrations in infected individuals ^17^, but still shows enhanced diagnostic performance when compared to the composite reference standards^18^. *Mtb*-ccfDNA is released during apoptosis and lysis of *Mtb* ^19,20^, and accumulates in the bloodstream during tissue perfusion making it possible to detect it in plasma. The emergence of the ddPCR technology has paved way for the detection of very low copies of target with high levels of sensitivity. The high level of reported sensitivity of the ddPCR technology and enhanced diagnostic utility of *Mtb* ccf DNA detection in the diagnosis of TB prompted us to explore the use of ddPCR to detect circulating *Mtb-*ccf DNA as a biomarker for early identification of asymptomatic *Mtb-*infected individuals harboring low numbers of live *Mtb*, and therefore at high risk of progression to active TB disease sooner or later.

Our study for the first time demonstrates the presence of *Mtb*-derived ccfDNA in the blood of individuals with a high risk of progression to active TB disease. Using the ddPCR technology, we were able to detect circulating *Mtb* DNA in 79·0% of progressors, as early as 6 months prior to TB breakdown, highlighting the great promise held by this method as a predictive test for early identification of individuals with a high risk for developing infectious TB disease. In the case of EPTB, we could detect ccf DNA in 100·0% of Progressors even at six months prior to TB diagnosis. Very interestingly, we could detect ccf DNA in about 50·0% of at-risk individuals even as early as 18 months (19-28 months) prior to actual TB breakdown, which we believe is a breakthrough observation. The diagnostic performance of this test clearly meets the Target Product Profile (TPP) framed by the WHO for tests that predict TB progression ^21^.

We further analysed the performance of the ddPCR test in establishing the diagnosis of TB in various groups of Progressors/Incident TB cases as defined in Table 1. We could detect *Mtb-* ccfDNA in the plasma of 9/11 clinically diagnosed possible TB cases and 10/11 subclinical TB cases rendering a remarkable sensitivity of 81·8% and 90·9% respectively, highlighting the usefulness of the technique in providing an accurate diagnosis in instances where the diagnosis cannot be established with confidence using the currently available routine diagnostics.

In many peripheral set-ups, due to various existing challenges, clinicians resort to empirically starting patients on anti-TB treatment without a confirmed diagnosis of tuberculosis. For instance, in a study done in Uganda, despite negative Xpert results, clinical diagnosis and empirical treatment detected two extra TB cases confirmed by culture, although this came at the cost of treating 18 patients with negative TB cultures unnecessarily^22^. In a high TB prevalence country like India, apart from diagnostic limitations, other factors like risk of losing a patient and the pressure to provide symptomatic relief drive clinicians to start them on empirical treatment ^23^. Even more precarious is the missing out of potential TB cases because of lack of confirmed evidence of the disease. This crucial scenario calls for an accurate, highly sensitive diagnostic test that can directly detect *Mtb* in easily accessible samples such as plasma or urine. Detection of IS6110 and IS1081 sequences of *Mtb* in *Mtb-*ccfDNA can be a highly valuable test to help clinicians take a clear diagnostic call in the scenario of non-availability of a highly sensitive microbiological and molecular test. Furthermore, the method appears to hold great potential for the early detection of EPTB, which has always been challenging given the insidious clinical presentation and the difficulty in collecting an appropriate sample ^24^. The standardised ddPCR protocol detected *Mtb* DNA in all EPTB patients giving a sensitivity of 100·0% which is highly encouraging.

Our study demonstrates the superior diagnostic capability of ddPCR targeting two *Mtb-*specific targets at the instance of TB breakout as compared to the currently available routine TB diagnostic tests, giving a sensitivity of 87·0% as against 76·1% for the currently employed diagnostic tests put together. Previous studies on detection of ccf DNA in plasma samples of TB patients had yielded much lower detection rates of 42·6% and 65·0% ^11,17^, though higher levels of sensitivity have been reported with sputum samples ^25,26^.

The LOD of ddPCR for detecting *Mtb-*ccfDNA was as low as 1 copy/20μL of plasma, which makes the assay highly sensitive. We eliminated the likelihood of false positive results by running multiple negative and positive controls in every assay to ensure the detection of true positives only. Various studies have reported that pre-analytical factors like blood collection, processing and ccfDNA extraction can interfere with the performance of the ddPCR assay ^27^. However, all our analysis was performed on retrospectively collected and stored plasma samples emphasizing that appropriately collected and properly stored plasma can very well be used for this test. We used the bead-based extraction method for ccfDNA extraction since the yield was much better than the column-based extraction method. The low abundance of ccfDNA in blood however necessitates a larger input volume of plasma for the assay. Through standardization experiments we found that 1 mL of plasma was suitable for optimal detection. While presenting the encouraging findings of our study, we are further tempted to speculate that multiplexing the assay with more *Mtb* targets in addition to IS6110 and IS1081, such as CFP10 and RD1, and the use of fresh samples can further improve the sensitivity of detection of the ddPCR assay.

While this test would require skilled personnel and a laboratory set-up, the test would still be useful in resource limited settings, as the estimated cost per test is nearly the same as that of the other nucleic acid-based TB tests. While the limitation of the present study is the small sample size, its strength lies in the use of well-preserved samples obtained from well characterized cohorts of HHCs who were systematically followed up for outcome evaluation. Validation of the IS6110-IS1081 ddPCR assay in multicentric cohorts for early identification of high risk individuals, as well as in diagnostically challenging cases would help authenticate the utility of this test in the field settings.

## Data Availability

All data produced in the present work are contained in the manuscript

