## Supplementary Figure 1 for "Spotting the Silent Threat: Early Detection of Subclinical Tuberculosis in High-Risk Individuals"

**Supplementary Table 1. Primer and probe sequences of IS6110 and IS1081 targets**

| Primers and Probes | Sequence 5'→3' |
|---|---|
| IS6110 Forward | AGCGCCGCTTCGGACCACCAG |
| IS6110 Reverse | AGGCGTCGGTGACAAAGGCCACGTA |
| IS6110 Probe | FAM-CGGCTGTGGGTAGCAGACCTCACC-BHQ1 |
| IS1081 Forward | CAGCCCGACGCCGAATCAGTTGTT |
| IS1081 Reverse | GGTGCGGGCGGTGTCGAGGTG |
| IS1081 Probe | FAM-CGCAGCGGTACTCGACGCTCTGACCGACAAGCTGCG-BHQ1 |

**Supplementary Figure 1. Participant selection from the study cohort**

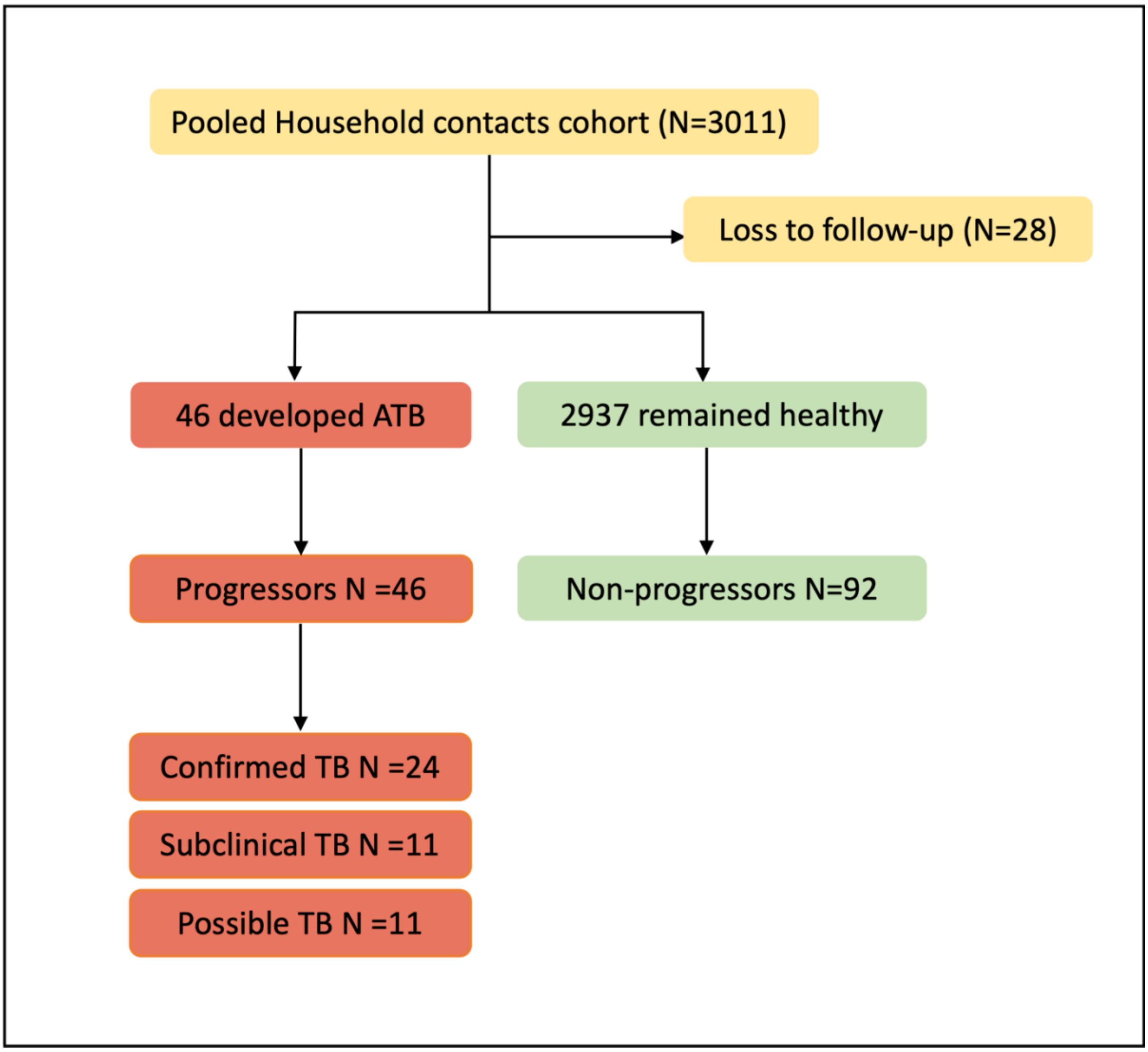

**Supplementary Figure 2. Representative amplitude plot and event counts of the ddPCR assay**

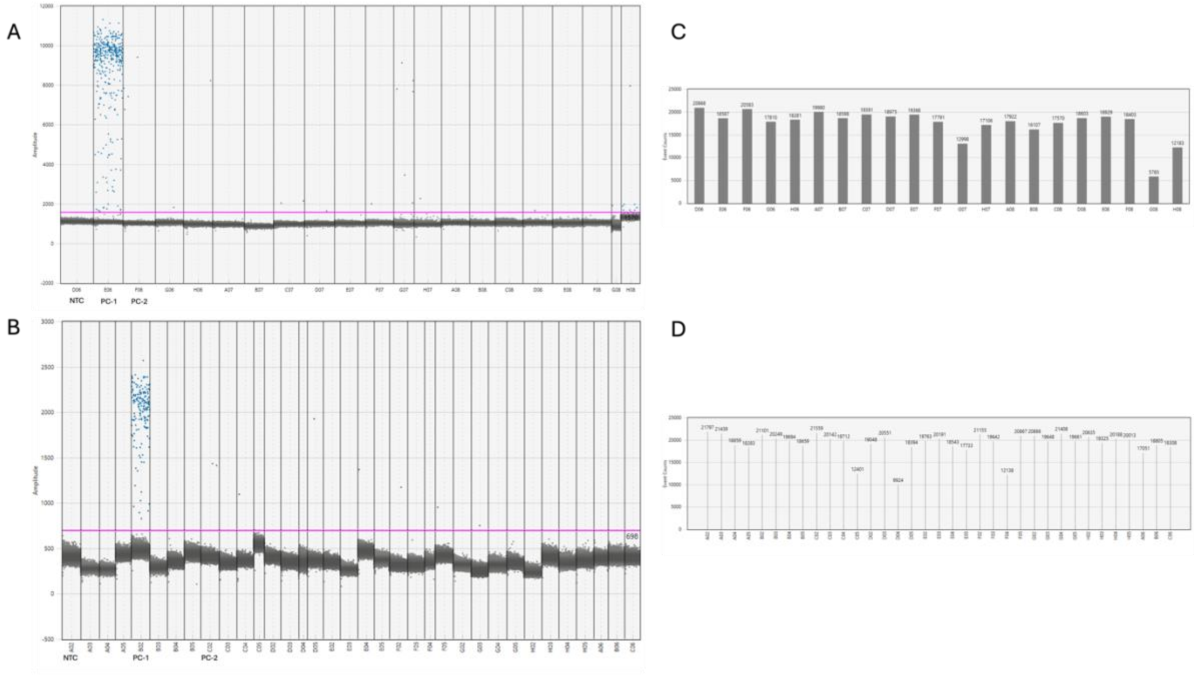


*(A) Amplitude plot of IS6110 assay (B) Amplitude plot of IS1081 assay (C) Event counts of IS6110 assay (D) Event counts of IS1081 assay*

*NTC: No template control; PC-1: Positive control H37Rv (0.003ng), PC-2: Positive control H37Rv (3fg)*
